# Low-dose interleukin-2 in birch pollen allergy: a phase-2 randomized double-blind placebo-controlled trial

**DOI:** 10.1101/2024.09.04.24312986

**Authors:** M Rosenzwajg, A Gherasim, F Dietsch, M. Beck, N Domis, R Lorenzon, Yannick Chantran, B Bellier, E Vicaut, A Soria, F De Blay, D Klatzmann

**Author notes:** Corresponding Author: Prof. D. Klatzmann, Hôpital Pitié-Salpêtrière, Clinical Investigation Center for Biotherapies and Immunology-Immunopathology-Immunotherapy department, Hôpital Pitié-Salpêtrière, 83 Bd de l’Hôpital, F-75013, Paris, France.

## Abstract

**Background:** Regulatory T cells (Tregs) are pivotal in immune tolerance to allergens. Low-dose IL-2 (IL-2_LD_) expands and activates Tregs. We assessed IL-2_LD_ efficacy for controlling clinical responses to allergen exposures.

**Methods:** RHINIL-2 was a phase-2a single-centre, randomised, double-blind, placebo-controlled proof-of-concept study. Twenty-four patients with allergic rhinitis to birch pollen (BP) were included, 66% having concomitant asthma. All had IgE and positive skin prick tests to BP at inclusion, and a total nasal symptom score (TNSS) ≥5 following a 4-hour nasal exposure to BP in an environmental-exposure-chamber (EEC). Patients received 1 MUI/day of IL-2 or Placebo for 5 days, followed by weekly injections for 4 weeks. Clinical responses to subsequent BP exposures in the EEC were evaluated using TNSS, the rhinitis visual analogue scale (VAS) and spirometry. The primary efficacy endpoint was the difference in TNSS area under the curve between inclusion and day 40 (TNSSΔAUC). This trial is registered with ClinicalTrials.gov (NCT02424396).

**Outcomes:** IL-2_LD_ treatment induced a significant expansion and activation of Tregs. The TNSSΔAUC in the ILT-101 and Placebo groups was non significantly different (-8.03 vs -4.76, p=0.32). TNSS and VAS AUCs were significantly reduced from baseline to day 40 in the ILT-101 group only (p=0.04 and p=0.01 respectively). The ratio of forced expiratory volume in 1 second / forced vital capacity (FEV_1P_) and the forced mid-expiratory flow (FEF_25-75%_) showed significant improvement in the ILT-101 vs Placebo groups at day 40 (p=0.04 and 0.04 respectively). There was a significant increase in eosinophils during treatment and no severe treatment-related adverse events.

**Interpretation:** IL-2_LD_ is well-tolerated in allergic patients, including in those with asthma. Although there was a trend towards a reduction in nasal scores, the primary endpoint was not reached in this small study. However, the short treatment duration used in this study cannot have effects on IgE levels given their half-life. Thus the limited efficacy observed suggest that Tregs mitigate allergic reactions and represent novel targets for the treatment of allergy.

**Funding:** Assistance Publique-Hôpitaux de Paris, ILTOO pharma, Agence Nationale de la Recherche

**Research in the context:** Allergic rhinitis (AR) is a common inflammatory disorder of the nasal mucosa, affecting millions worldwide, and often associated with asthma. Its management encompasses various strategies targeting symptom relief, such as antihistamines, corticosteroids and leukotriene receptor antagonists. Biologics targeting specific pathways, such as IgE, IL-4, IL-5, and IL-13, are in development. Curative treatment such as allergen-specific immunotherapy (AIT) for re-establishing tolerance to the allergen have limited efficacy. Despites its often moderate severity, AR can profoundly affect the quality of life and remains an unmet medical need.

Treg have a clear and direct role in preventing allergy, as exemplified by the fact that their complete deficiency in the IPEX syndrome leads to allergy. The role of Treg in mitigating an existing allergy is less clearly established. It mainly comes from the observation that successful allergen-specific immunotherapy (AIT) is associated with the induction of Tregs. Of note, both a Treg defect and a positive impact of Tregs during AIT have been described for allergic rhinitis patients. Collectively, these results highlight that strategies to increase Treg numbers and/or fitness might be beneficial in the treatment of allergic rhinitis.

**Evidence before this study:** Treg-targeted therapies have not yet been evaluated in humans with allergy. That IL-2_LD_ has not yet been evaluated is possibly because it triggers eosinophilia. This is due to the stimulation of innate lymphoid cells type 2 (ILC2), which express the high affinity receptor for IL-2 and produce IL-5 upon IL-2 activation, which in turn triggers the eosinophilia. Eosinophils are critical mediators in allergic responses, contributing to inflammation and tissue damage. When exposed to allergens, eosinophils release molecules, including histamines, leukotrienes, and cytokines, which contribute to tissue inflammation and allergy symptoms. Eosinophils are particularly implicated in asthma and allergic rhinitis, in which they contribute to airway hyperresponsiveness, mucus production, and remodelling. IL-5 is a key cytokine for eosinophils and monoclonal antibodies against IL-5 are currently developed. Of note, the IL-2_LD_-triggered IL-5-induced eosinophilia has not yet been associated with severe side effects, even in patients who have received daily IL-2_LD_ injections for years and had persistent eosinophilia.

**The added value of this study:** This is to our knowledge the first study of a Treg-targeted therapy in allergy, and of IL-2LD in allergy. It shows that, as expected, IL-2LD can directly stimulate Tregs and indirectly eosinophils in patients with allergies. The eosinophilia went up to twice the normal value and had no clinical significance, including in patients with asthma. The IL-2LD safety profile in this double-blind placebo control study relieves the concerns of using it in allergy, and thus license its further clinical investigation, including in asthma. Although there was a trend towards a reduction in nasal scores, the primary endpoint was not reached in this small study. However, the short treatment duration used in this study cannot have effects on IgE levels given their half-life. Thus, the limited efficacy observed suggest that Tregs mitigate allergic reactions and represent novel targets for the treatment of allergy that warrants further clinical investigation in larger studies. Our study also highlights the value of EEC for studying a novel treatment of allergy.

**Implications of all the available evidence:** The possible improvements in the clinical response to an allergen challenge were obtained after a short treatment that stimulated Treg fitness but could not have any effects on the effector mechanisms of allergy. Thus, as they showed that Treg could mitigate ongoing allergic response, Treg represents a novel target in allergy. This opens the door for combination therapies, notably with molecules targeting the effector immune responses and with allergen-specific therapies aimed at re-establishing tolerance to the allergen.

## Introduction

Allergic diseases such as rhinitis, asthma, food allergy, atopic dermatitis, and other skin complaints are among the commonest chronic disorders, affecting up to 15-30% of the population. Their prevalence over the last 20-30 years has been increasing dramatically. There is still no curative treatment for allergy, and novel therapies are needed.

The recent discovery of regulatory T cells (Tregs), which regulate all immune responses, has dramatically changed our understanding of the pathophysiology of allergic diseases and opened new therapeutic perspectives. Treg impairment is associated with loss of tolerance, autoimmunity, and also allergy^1^. Indeed, allergy is one of the clinical manifestations of patients with IPEX syndrome, a Treg deficit caused by mutations in FOXP3^2^. Dysfunctional Tregs have been identified in allergic individuals^3–5^ and children at high risk of allergic diseases^6^. The protective role of Tregs in allergy is further supported by the fact that successful allergen-specific immunotherapy (AIT) is associated with the induction of Tregs that were shown to suppress different effector mechanisms associated with allergy^7,8,9^. Several mechanisms could contribute to the Treg-mediated suppression of allergic diseases^10^: (i) reduced priming of allergen-specific Th2-type T cells ^3,11^ and B cells^12^, (ii) switch from allergen-specific IgE to IgG4 production^13^ and (iii) downregulation of mast cell degranulation via inhibition of calcium influx^14^ or expression of FcεRI^15^. Collectively, these results highlight that allergic diseases result from an inappropriate balance between allergen-activated effector Th2 cells and Tregs. As a result, Treg-based therapies have the potential to re-equilibrate this balance and improve or even prevent allergic diseases^16^.

Importantly, both a Treg defect and a positive impact of Tregs during AIT have been described for allergic rhinitis patients. The proportion of Tregs is significantly reduced in children with allergic rhinitis compared to healthy controls^17,18^. More recently, it was shown that Notch signalling downregulates Foxp3 expression and likewise inhibits the differentiation of Treg cells which consequently promotes the development of allergic rhinitis^19^. Pollen-specific immunotherapy has been shown effective for IgE-mediated seasonal allergic rhinitis, involving the recruitment and activation of Tregs^20^. Successful grass pollen immunotherapy was associated with markedly higher numbers of Tregs in the nasal mucosa^21^. Collectively, these results highlight that strategies to increase Treg numbers and or fitness might be beneficial in the treatment of allergic rhinitis.

This could be achieved with low-dose IL-2 (IL-2_LD_) which specifically expands and activates Tregs, blocks the development of naïve CD4 cells in pro-inflammatory Th17 cells^22^, and blocks the development of naïve CD4 cells in follicular helper T cells (Tfh)^22^ that are important for the affinity maturation of B cells and the development of auto-antibodies. In line with these properties, we recently demonstrated that IL-2_LD_ can specifically expand/activate Tregs in vivo and can control allergic reactions in two models of food and respiratory allergy in mice^23^. Similarly, a complex of IL-2 bound to an anti-IL-2 antibody, which has been shown to also induce Treg expansion, suppressed allergic airway disease in mice^24,25^.

There is thus a strong rationale for evaluating IL-2_LD_ as a treatment for allergy. However, IL-2 is known to also stimulate ILC2 cells to produce IL-5, in turn inducing eosinophilia^26^. While the role of eosinophils in the pathophysiology or allergy is debated^27^, an accumulation of eosinophils in the airways is characteristic of allergic rhinitis and asthma. IL-2_LD_ effect on eosinophils is dose-dependent and only moderate at the dose used, and there are so far no indication of any allergic severe adverse events in patients treated with IL-2_LD_, even in patients with permanent eosinophilia during treatments for over 5 years^28,29^. Nevertheless, to our knowledge, IL-2_LD_ has not been used to treat allergy and there is a theoretical risk that IL-2_LD_ could aggravate allergy. Therefore, we conducted a proof-of-concept clinical trial of IL-2_LD_ in patients with allergic rhinitis to birch pollen, as an initial step before evaluation in other more severe forms of allergy.

Rhinil-2 trial evaluates IL-2_LD_ in patients with allergic rhinitis to birch pollen, within a randomized, placebo-controlled, double-blind, phase IIa clinical trial (NCT03776643). To evaluate allergic rhinitis in a safe and standardized manner, we used ALYATEC Environmental Exposure Chamber (EEC). This chamber was previously validated with 3 different allergens in rhinitis and conjunctivitis related to birch pollen and cat and mite asthma. This allowed for monitoring of the concentration of airborne allergens as well as the particle size (number and aerodynamic diameter) to maintain high reproducibility between allergen exposures and guarantee both the quality of clinical data and the safety of patients^30–32^. The primary objective of the study was the evaluation of the efficacy of ILT-101 (IL-2_LD_) to reduce the nasal response during allergen exposures as compared to placebo.

## METHODS

### Study design, participants and treatment

#### Design

RHINIL-2 was a phase IIa monocentric randomized, double-blind parallel-group, placebo-controlled (1 active / 1 placebo) proof-of-concept trial evaluating the tolerance and the efficacy of IL-2_LD_ in adult patients affected by allergic rhinitis to birch pollen. The primary endpoint was the change in the 0 to 4 hours TNSS Area under the curve (AUC 0-4h) at day 40 compared to baseline, between the treated and placebo groups. Clinical secondary endpoints included changes from baseline in TNSS on Day 8 and a visual analogue scale (VAS) and spirometry for forced expiratory volume in 1 second (FEV_1_ and the forced mid-expiratory flow (FEF_25-75%_). Biological secondary endpoints included changes in Tregs and eosinophils. Safety was assessed throughout the study (**Figure S1**).

The study was approved by the institutional review board CPP Sud-Ouest et Outre-Mer II and performed in accordance with the Declaration of Helsinki and good clinical practices. RHINIL-2 is registered as “*<u>Safety and Efficacy of Low-dose IL-2 in Birch Pollen Allergy</u>*” with EUDRACT number 2017-002907-90 and with ClinicalTrials number NCT03776643.

#### Patients

Patients were recruited in Strasbourg, France, and were randomised, treated, and followed up at the Alyatec Center in Strasbourg Civil Hospital. Patients were eligible if they were aged 18-55 years with a history of allergic rhinitis to birch pollen, with or without associated asthma. The main exclusion criteria were having a history of anaphylactic reactions, severe hematological disorders, vital organ failure, cancer and active infections (supplementary table S1). Among 60 eligible patients for the study, 27 were selected, and 24 were randomized (**Figure 1**). Written informed consent was obtained from all participants before enrolment in the study.

**Figure 1:**
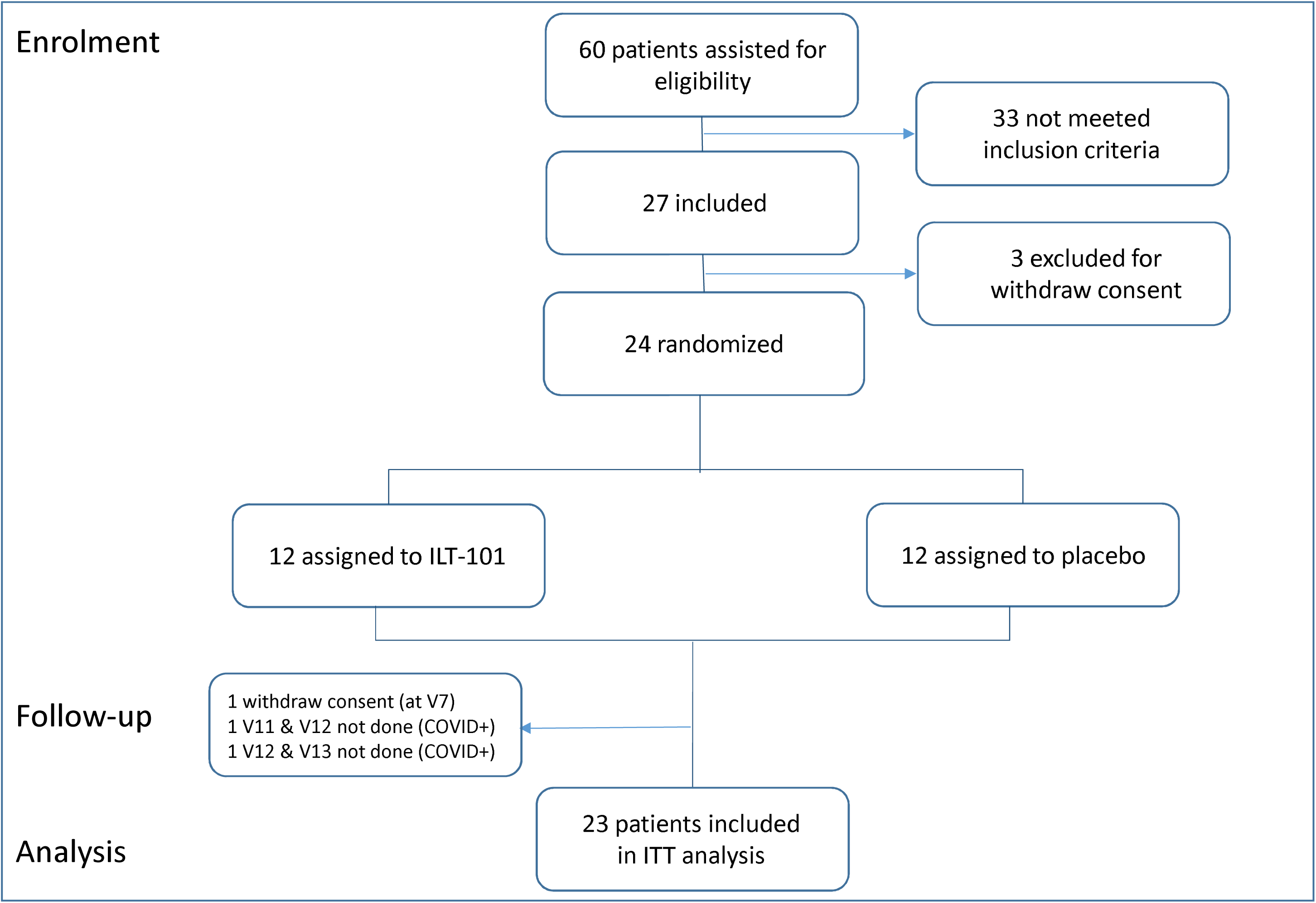
Trial Profile.

#### Treatment

ILTOO Pharma provided the drug (IL-2_LD_: ILT-101®) and placebo (mannitol and sodium dodecyl sulfate) in the same packaging to the Alyatec pharmacy. ILT-101® is a formulation of adesleukin that is packaged in ready-to-use vials of 1.2 MIU. Treatment administration consisted in a first course of daily injections for five days (the induction course), followed by a single injection every week for 4 weeks (the maintenance course; from day 15 to day36) (**Figure S1**).

### Randomization and masking

The computer-generated randomization list was managed by an independent statistician. Patients were randomized in a 1:1 ratio to ILT-101® or placebo. Patients were given a study number according to the patient’s order of entry into the study. All personnel in contact with the patients (research nurses, investigators, clinical research assistants) and assessing outcomes (including laboratory ones) were masked to group assignment until the end of study analyses. Syringes containing placebo and IL-2LD had the same appearance and were labelled according to good manufacturing practice for traceability and accountability purposes.

### Patients monitoring

Visits occurred i) during the screening period, the patient underwent the required tests and first exposure to EEC (baseline) to confirm eligibility, ii) during the treatment period, from day 1 to day 5 and on days 8, 15, 22, 29, and 36, and iii) during the follow-up period at day 40 and at day 70. Blood samples for immunological tests were obtained at screening, and on days 1, 8 and 40. Safety was assessed throughout the study.

### Environmental Exposure Chamber (EEC)

ALYATEC EEC is a 65 m^2^ International Standards Organization (ISO) 8 clean room (Standard 14644-1) with continuous air circulation, containing 20 seats to accommodate up to 20 patients at 1 time, and with an anteroom and exit room. The EEC contains 10 particle counters spread equally throughout the room, and 5 glass fibber filters adjacent to seats to be used for Bet v 1 allergen testing by ELISA. Standardized, commercially available birch pollen extract prepared by a pharmacist is nebulized into the EEU through a specialized system that maintains consistent birch allergen particle counts and Bet v 1 ELISA dosing approximating 60 ng/m^3^ across the 20 seat positions of the EEC. Allergen particle plateau fall below the limit of detection within minutes of stopping the nebulization and Bet v 1 measurements are below the limit of quantification outside of the EEC (e.g., in the observation area and in the rest rooms). Other allergens have been tested in the EEC, observation room, and the rest rooms and are below the limit of quantification (e.g., house dust mite, cat), suggesting that there is no cross-contamination of allergens. During each exposure, the patients wear protective suits (TS Plus Microgard 2000) to ensure that no contaminant enters or exits the EEU. The cleaning procedure after each EEU exposure session includes the following: rinsing of the nebulization system, disinfectant of the floor, walls, the armchairs, and of all the small material stored in the EEU (Ecowipes, THX medical). Additionally, air qualification is performed by an independent company (Air Qualif) 4 times a year to test for the absence of bacteria and molds in the EEU. This facility enables the provision of a homogenous distribution of allergens for each participant. The airflow system, environmental conditions, and airborne particles are continuously monitored. Communication between participants inside the EEC and with medical supervisors was enabled using wireless telecommunication.

The same batch of birch allergen extract (Allergopharma®) was used forall exposures. Participants entered the EEC once the plateau of airborne allergen was reached.All exposures lasted up to 4 h. Homogeneous allergen distribution was verified using particle counters, as previously described ^30,32,31^. After each exposure, the Bet v 1 concentration was determined using ELISA (Indoor Biotechnologies®, Charlottesville, VA, USA) by collecting allergen on glass fiber filters located next to the participants’ chairs. This EEC used in the current study was previously validated for use in birch allergen, where standardized and reproducible allergen levels were achieved on repeated exposures^32^.

### Nasal allergen challenge evaluation

The total nasal symptom score (TNSS) is a validated questionnaire used to evaluate nasal responses in allergic rhinitis studies^32^. This score is the sum of scores for nasal congestion, sneezing, nasal itching, and rhinorrhoea at each time-point, using a four-point scale (0–3), where 0 indicates no symptoms, 1 indicates mild symptoms that are easily tolerated, 2 indicates moderate awareness of symptoms that are bothersome but tolerable, and 3 indicates severe symptoms that are difficult to tolerate and interfere with activity. TNSS was evaluated with a maximum score of 12 and considered positive if the scores differed by at least 5 points from baseline^32^. Changes in nasal response determined by TNSS were expressed as area under the curve (AUC) for measures assessed every 20 min during the 4 hours of exposure.

Participants also self-evaluated the severity of rhinitis using a visual analogue scale (VAS) for rhinitis. VAS is a semiquantitative subjective means of evaluating nasal symptoms on a scale of 0–100 mm, where 0 indicates no symptoms. The threshold was at least 23 mm, as previously described^33^. Participants self-evaluated their severity of rhinitis by positioning the cursor of the scale on an electronic tablet.

### Bronchial assessment by spirometry

To promote safety, spirometry was performed during allergen challenges (Spiro Bank II, MIR®, France). Forced expiratory volume in 1 second (FEV_1_) and other parameters (FVC, FEV_1_P, FEF_25-75%_ and PEF) were assessed every 30 min during the 4h birch exposure, with supplementary assessments if asthma symptoms developed. A 20% drop in FEV_1_ during a 4-h exposure was defined as an early asthma response (EAR). The same protocol has already been used in this EEC for validation in asthmatic patients to cat and house dust mites^34^.

### Immunomonitoring

All blood samples were acquired immediately before the administration of IL-2_LD_. All the immunomonitoring procedures (flow cytometry and quantification/analysis of cytokine and levels and Basophile activation test) were performed as previously described^35–38^ and are described in the Supplementary methods.

### Outcomes

The primary outcome was the change in nasal response determined by TNSS, expressed as area under the curve (AUC), from baseline to day 40, between ILT-101 and placebo. Clinical secondary outcomes were changes in TNSS AUC and VAS AUC from baseline on day 8. Biological secondary outcomes were changes from baseline (absolute and relative) on days 8 and 40 (i) in Tregs and eosinophils, and (ii) in allergen-specific IgE and IgG4 dosages.

#### Statistical analysis

All analyses were performed in all randomized patients (intention to treat). The global between-group differences in the time-dependent profile of changes in clinical parameters and Tregs or other immunological parameters was analysed throughout the treatment period by non-parametric ANCOVA for repeated measurements, using baseline values as covariable (Conover method). Data are presented as mean ± SD or average (range). Normal distribution of each set of variables was analysed by D’Agostino & Pearson omnibus normality test. Student’s t-test for normally distributed data and non-parametric Mann– Whitney were used to compare variables as appropriate. Paired data comparing day 8 or day 40 parameters to baseline value were analysed using Student’s t-test for normally distributed data and paired Wilcoxon signed-rank test for nonparametric. A p value < 0.05 was considered significant. Statistical analyses were performed using GraphPad Prism version 10.0 (GraphPad Software, San Diego, CA, USA).

## Results

### Study Design and Patients

Sixty patients were assessed for study eligibility between February 2019 to August 2022. Thirty-three patients did not meet the inclusion criteria, three patients withdrew consent before randomization and twenty-four patients were randomized in two parallel groups (1 ILT-101/1 placebo). One patient withdrew consent on day 5 and two patients did not attend because they tested positive for COVID-19. Twenty-three patients were included in the intention-to-treat analysis for primary and secondary endpoints (**Figure 1**).

Baseline demographics and disease profiles are described in **Table 1** . All patients included had concomitant allergies; they had a mean age of 33 years and a mean disease duration of about 21 years. More than half of the patients in the two groups had a history of mild to moderate asthma. Demographic characteristics and disease-related clinical and biological values were balanced between the ILT-101 and placebo groups (**Table 1**).

**Table 1:** Demographics characteristics.

|  | ILT-101<br>N=12 | Placebo<br>N=12 | Total population<br>N=24 |
| --- | --- | --- | --- |
| **Age, years**<br>mean (± SD)<br>range | 32.8 (± 9.5)<br>20, 49 | 33.2 (± 9.4)<br>22, 52 | 33.0 (± 9.3)<br>20, 52 |
| **BMI, kg/m<sup>2</sup>**<br>mean (± SD)<br>range | 23.8 (± 3.9)<br>20.5, 35.0 | 21.9 (± 2.4)<br>18.8, 26.7 | 22.8 (± 3.3)<br>18.8, 35.0 |
| **Male/Female**<br>N<br>(%) | 5/7<br>(41.7%/58.3%) | 3/9<br>(25.0%/75.0%) | 8/16<br>(33.3%/66.7%) |
| **Disease duration, years**<br>mean (± SD)<br>range | 19.44 (± 9.1)<br>6.27, 37.26 | 22.46 (± 11.24)<br>6.28, 46.98 | 20.95 (± 10.12)<br>6.27, 46.98 |
| **Concomitant allergic diseases**<br>n (%) | 12 (100.0%) | 12 (100.0%) | 24 (100.0%) |
| **History of asthma**<br>n (%) | 9 (75.0%) | 7 (58.3%) | 16 (66.7%) |
| **TNSS**<br>mean (± SD)<br>range | 1.3 (± 0.9)<br>0, 3.0 | 1.1 (± 1.1)<br>0, 3.0 | 1.2 (± 1.0)<br>0, 3.0 |
| **Nasal Response Intensity, VAS**<br>mean (± SD)<br>range | 1.0 ± 0.9<br>2.0 0.0, 3.1 | 0.7 ± 0.7<br>0.0, 1.7 | 0.8 ± 0.8<br>0.0, 3.1 |
| **Nasal Response Intensity (VAS)**<br>**AUC<sub>0-4h</sub> during exposure period**<br>mean (± SD)<br>range | 14.29 ± 7.86<br>4.83, 26.90 | 14.27 ± 7.01<br>6.57, 27.33 | 14.28 ± 7.28<br>4.83, 27.33 |
| **FEV 25-75**<br>mean (± SD)<br>range | 3.70 (± 1.15)<br>1.70, 5.90 | 3.95 (± 0.98)<br>2.66, 5.45 | 3.82 (± 1.05)<br>1.70, 5.90 |
| **FEV 1**<br>mean (± SD)<br>range | 3.62 (± 0.89)<br>2.23, 5.27 | 3.55 (± 0.65)<br>2.62, 4.67 | 3.58 (± 0.77)<br>2.23, 5.27 |
| **Birch allergen wheal diameter, mm**<br>mean (± SD)<br>range | 7.5 (± 1.1)<br>6.5, 10 | 7.1 (± 1.1)<br>6.0, 9.5 | 7.3 (± 1.1)<br>6.0, 10.0 |
| **Bet v 1 IgE, kU/L (± SD)**<br>mean (± SD)<br>range | 51.4 (± 61.2)<br>6.9, 225 | 22.2 (± 20.3)<br>4.1, 67.2 | 36.8 (± 47.0)<br>4.1, 220 |
| **Tregs cells, % among CD4<sup>+</sup>**<br>mean (± SD)<br>range | 8.9 (± 2.6)<br>4.9, 13.5 | 9.5 (± 2.6)<br>4.2, 14.0 | 9.2 (± 2.6)<br>4.2, 14.0 |
| **Tregs cells/mm<sup>3</sup>**<br>mean (± SD)<br>range | 79.6 (± 26.9)<br>49.6, 134.0 | 91.0 (± 32.4)<br>55.9, 163.1 | 85.3 (± 29.7)<br>49.6, 163.1 |
| **Eosinophils count Giga/L**<br>mean (± SD)<br>range | 0.27 (± 0.17)<br>0.11, 0.55 | 0.13 (± 0.08)<br>0.03, 0.22 | 0.2 (± 0.14)<br>0.03, 0.55 |

### Clinical response to IL-2_LD_

TNSS on day 40 did not show significant differences between active and placebo group as compared to baseline (**Figure 2A, Table S2**). A statistically significant decrease of the TNSS during exposure was observed on days 8 and 40 as compared to baseline for active treatment groups only. (**Figure 2B**). Improvement of rhinitis symptoms in the ILT-101 group assessed by the VAS showed the same pattern of response. Rhinitis VAS at baseline was comparable for the two groups (mean of 14.28 ± 7.28 for the overall populations) (**Table 1**), VAS AUC was significantly decreased on days 8 and 40 compared to baseline in ILT-101 patients, but not in the placebo group (**Figure 2C**).

**Figure 2:**
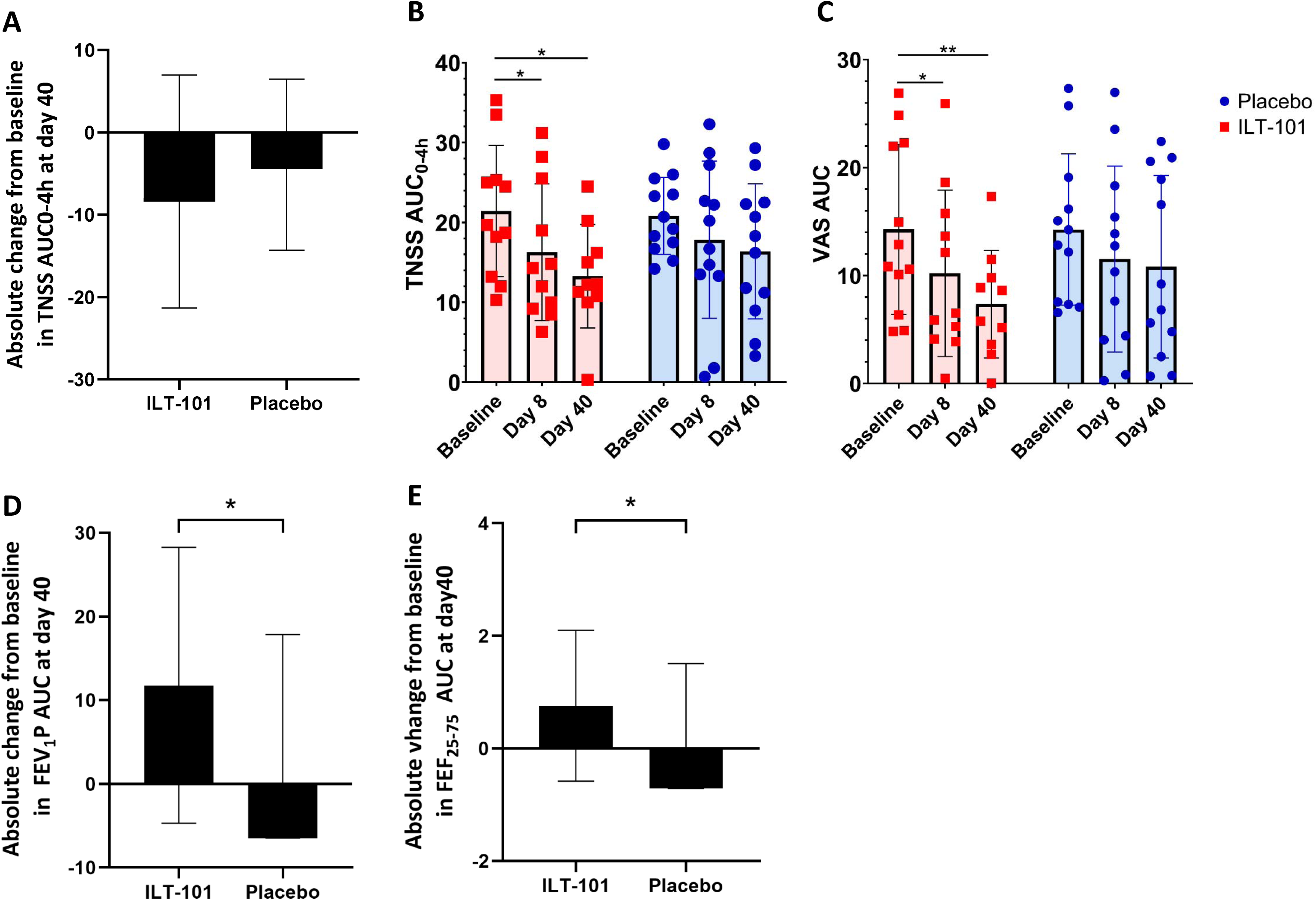
Clinical outcomes. **A)** Absolute change from baseline in nasal symptoms TNSS AUC at day40; **B-C)** Kinetics of TNSS and VAS AUC in ILT-101 (in red) and placebo (in blue); **D-E)** Absolute change in In FEF1P and FEF27-75 AUC at day 40 compared to baseline in ILT-101 and placebo groups. Data are represented as mean± sd and individual values. The global differences between the time-dependent profile of changes between treated and placebo patients throughout the treatment period was analysed by non-parametric ANCOVA for repeated measurements, using baseline values as covariable (Conover method). Comparisons of each parameter in treated patients at day 5 and day 40 were made using t-test or the Mann-Whitney test according to their normality distribution. (*p<0.05; **p<0.01; ***p<0.001).

Asthma response assessed by spirometry was also investigated every 20’ during exposures. It served to promote safety and offer further evaluation of the treatment effects. A decrease in forced expiratory volume in 1 second (FEV_1_) reveal an asthmatic response during the challenge. FEV_1_ was comparable in the two groups at baseline (**Table 1**). There was no decrease in this parameter during any of the allergen challenges, indicating no asthmatic response. Moreover, AUC FEV1P and FEF 25-75 were significantly augmented on day 40 in the ILT-101 treated patients compared to placebo-treated patients for whom AUC FEV_1_P and FEV _25-75%_ were decreased (**Figure 2D, 2E**).

### Safety

Treatment with IL-2_LD_ was generally well tolerated (**Table 2** ). No serious adverse events were reported in either group, placebo or ILT-101. Most of the adverse events reported were mild to moderate. The most frequent were injections site reactions, which occurred in 12 patients for 61 events (59%) in the ILT-101 arm and in 2 patients for 3 events (3%) in the placebo arm, followed by gastrointestinal symptoms [15 events 7 in patients (14%) and in 8 events in 4 patients (7.5%) respectively in ILT-101 and placebo arm], muco-cutaneous symptoms [9 events in 4 patients (9%) and 2 events in 1 patient (2%) respectively in ILT-101 and placebo arm], influenza-like syndrome [8 events in 4 patients (8%) and 3 events in 2 patients (3%) respectively in ILT-101 and placebo arm], asthenia [6 events in 4 patients (6%) and 2 events in 1 patient (2%) respectively in ILT-101 and placebo arm], headache [6 events in 5 patients (6%) and 2 events in 2 patients (2%) respectively in ILT-101 and placebo arm], and subclinical hypothyroidism [1 event in 1 patient (16%) and 2 events in 2 patients (2%) respectively in ILT-101 and placebo arm]. There were no unexpected adverse events reported. Of note, in line with the spirometry values, there was no clinical asthma detected during or after any of the allergen challenges.

**Table 2:** Safety.

|  | ILT-101<br>N° Event (N° Patients)<br>[% administrations] | Placebo<br>N° Event (N° Patients)<br>[% administrations] | Overall population<br>N° Event (N° Patients)<br>[% administrations] |
| --- | --- | --- | --- |
| Treatments administered | 103 (12) | 106 (12) | 209 (24) |
| Serious Adverse events (SAE) | 0 | 0 | 0 |
| Non Serious Adverse events (NSAE) | 202 | 61 | 263 |
| NSAE related to treatment | 115 [112%] | 23 [22%] | 138 [66%] |
| **The most common NSAE related to treatment** |  |  |  |
| Injection-site reaction | 61 (12) [59%] | 3 (2) [3%] | 64 (14) [31%] |
| Others AE | 54 (9) [53%] | 20 (6) [19%] | 74 (15) [35%] |
| Gastrointestinal symptoms | 15 (7) [15%] | 8 (4) [7.5%] | 22 (11) [10%] |
| Mucocutaneous symptoms | 9 (4) [9%] | 2 (1) [2%] | 11 (5) [5%] |
| Influenza-like syndrome | 8 (4) [8%] | 3 (2) [3%] | 11 (6) [5%] |
| Asthenia | 6 (4) [6%] | 2 (1) [2%] | 8 (5) [4%] |
| Headache | 6 (5) [6%] | 2 (2) [2%] | 8 (7) [4%] |
| Subclinical hypothyroidism | 1 (1) [1%] | 2 (2) [2%] | 3 (3) [1.4%] |

### Immune response

The mean ± SD baseline percentage of Tregs in patients was 8.9 ± 2.6 % of CD4+ T cells in the treated group and 9.5 ±2.6 in the placebo group (p=ns) (**Table 1** ). At day 8, an increase of Tregs to a mean of 13.5 ± 4.1 % was observed in the treated group compared to 9.8± 2.5 in the placebo group corresponding to a 1.52±0.21 and 1.04±0.15-fold increase respectively (p<0.0001) (**Figure 3A, Table S2** ). At day 40, Tregs increase was maintained in the treated group (12±3.7 = 1.33±0.17 fold change compared to 9.7 ± 2.3 =1.50±0.18 in the placebo group (p=0.0018) (**Table S2** ). The same observations were observed with Treg absolute numbers (**Figure 3B** ). As there were no effects of IL-2_LD_ on effector T cells, this translated into a significantly increased Treg/Teff ratio at day 8 and day 40 (**Figure 3C** ). At day 8, 72 hours after the last IL-2 injection, there was a highly significant increase of CD25 on Treg (**Figure 3E** ), a classical marker of IL-2-induced Treg activation^39,37,40,38^ associated with an increased concentration of soluble CD25 (**Figure 3F** ). Most of the other classical activation markers of Tregs had returned to baseline values at this time point (**Figure 3 D**).

**Figure 3:**
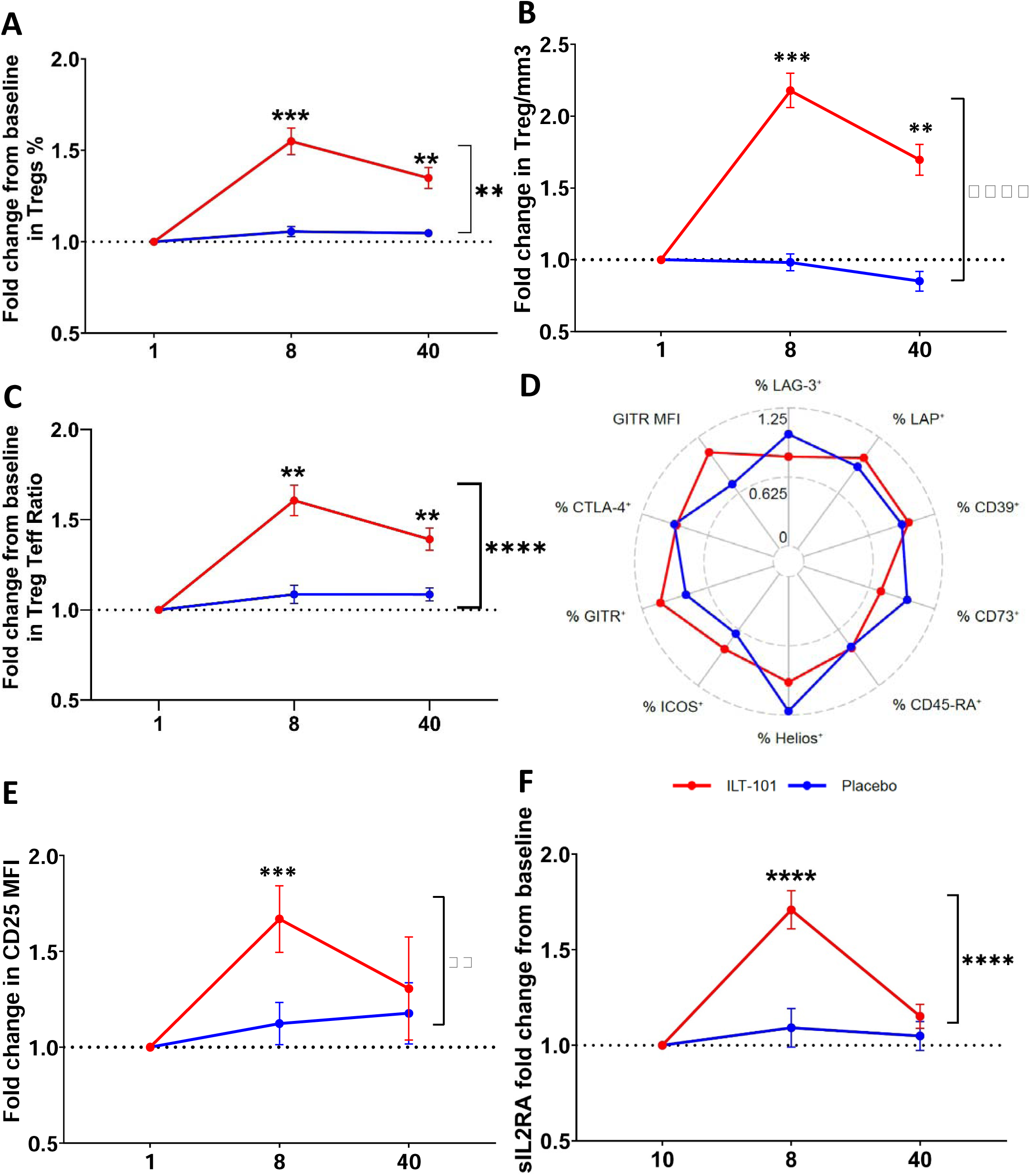
Changes in Tregs and Teffs. Treg cells were gated in CD4^+^ T cells and identified as CD25^hi^CD127^lo/-^Foxp3^+^cells. **(A-F)** Changes represented as mean ± sem by patient groups (IL-2 in red and Placebo in blue) from baseline to day 40, in **(A)** Tregs as percentages among CD4+ T cells;(**B**) Tregs absolute numbers; (**C**)Treg/Teff ratio; **(D)** Functional/activation markers expression by Treg; **(E)**. CD25 MFI in Tregs and (F) soluble IL2ra concentration. The global differences between the time-dependent profile of changes between treated and placebo patients throughout the treatment period was analysed by non-parametric ANCOVA for repeated measurements, using baseline values as covariable (Conover method). Comparisons of each parameter in treated patients at day 5 and day 40 were made using t-test or the Mann-Whitney test according to their normality distribution. (*p<0.05; **p<0.01; ***p<0.001).

### Other biological parameters

IL-2_LD_ led to a non-significant increase in the absolute number of eosinophils (**Figure 4A-B).** These remained within the normal values for most patients, reaching twice this value in two patients. There was no significant change in eosinophil’s activation markers such as DR, CD25, CD123, or CD44 (**Figure S2**)

**Figure 4:**
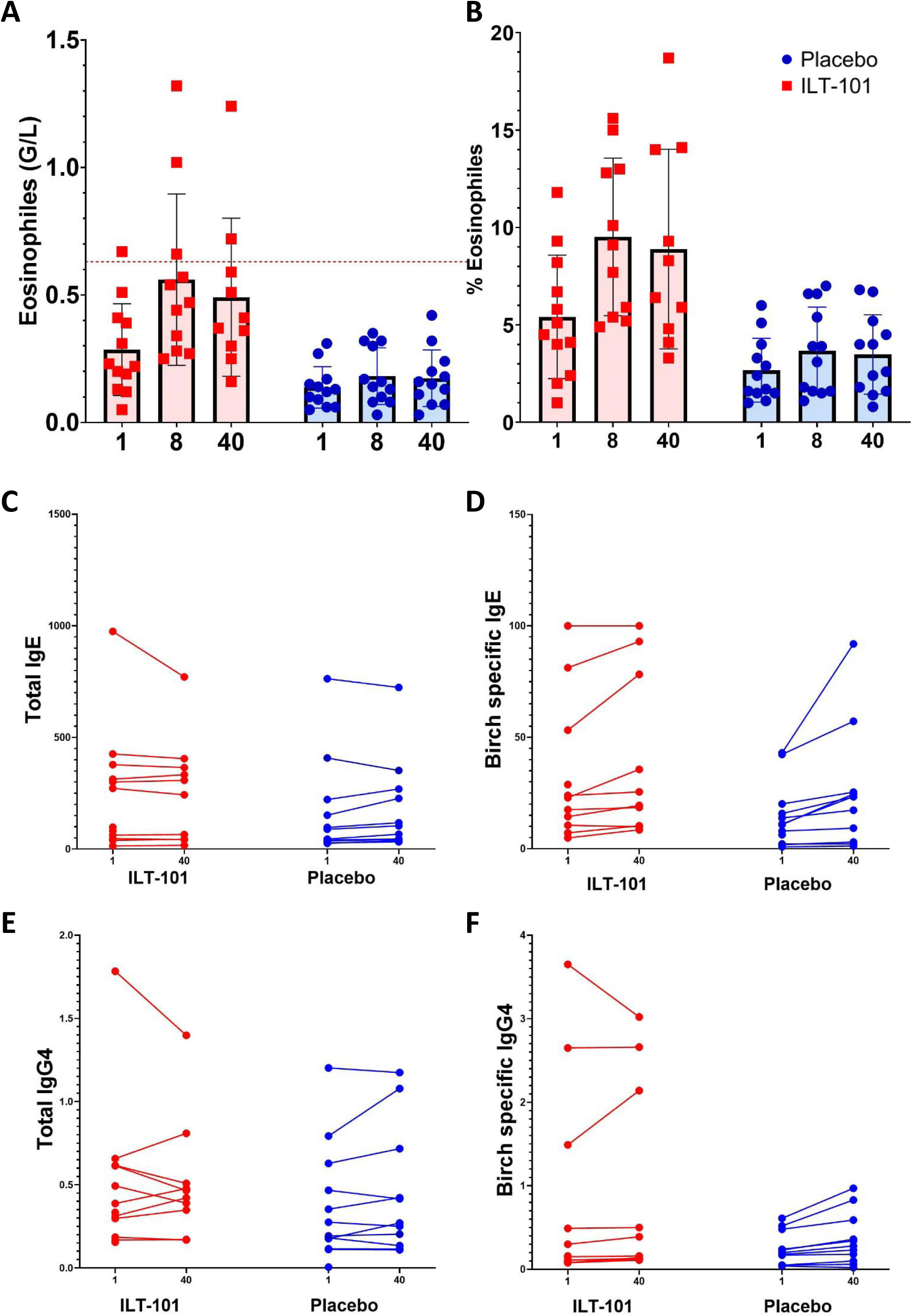
Changes in absolute and relative numbers of Eosinophils with IL-2_LD_. Eosinophil counts in Giga/L and percentage were measured all along the follow-up of patient. Results are shown as individual value and mean±sd for ILT-101trated group (in red) and in placebo (in vlue). Normal eosinophil counts in the local laboratory are 0.02 to 0.63 G/L and is showed as red dashed line.

As expected, considering the immunoglobulin half-life, there were no detectable changes in the concentration of total nor birch pollen-specific IgE as well as IgG4 (**Figure 4C-F**).

We also tested basophil degranulation after stimulation with serial dilution of birch allergen. We did not observe change over time in basophil allergen threshold sensibility and EC50 in ILT-101 nor placebo patients (**Figure 5A-C**).

**Figure 5:**
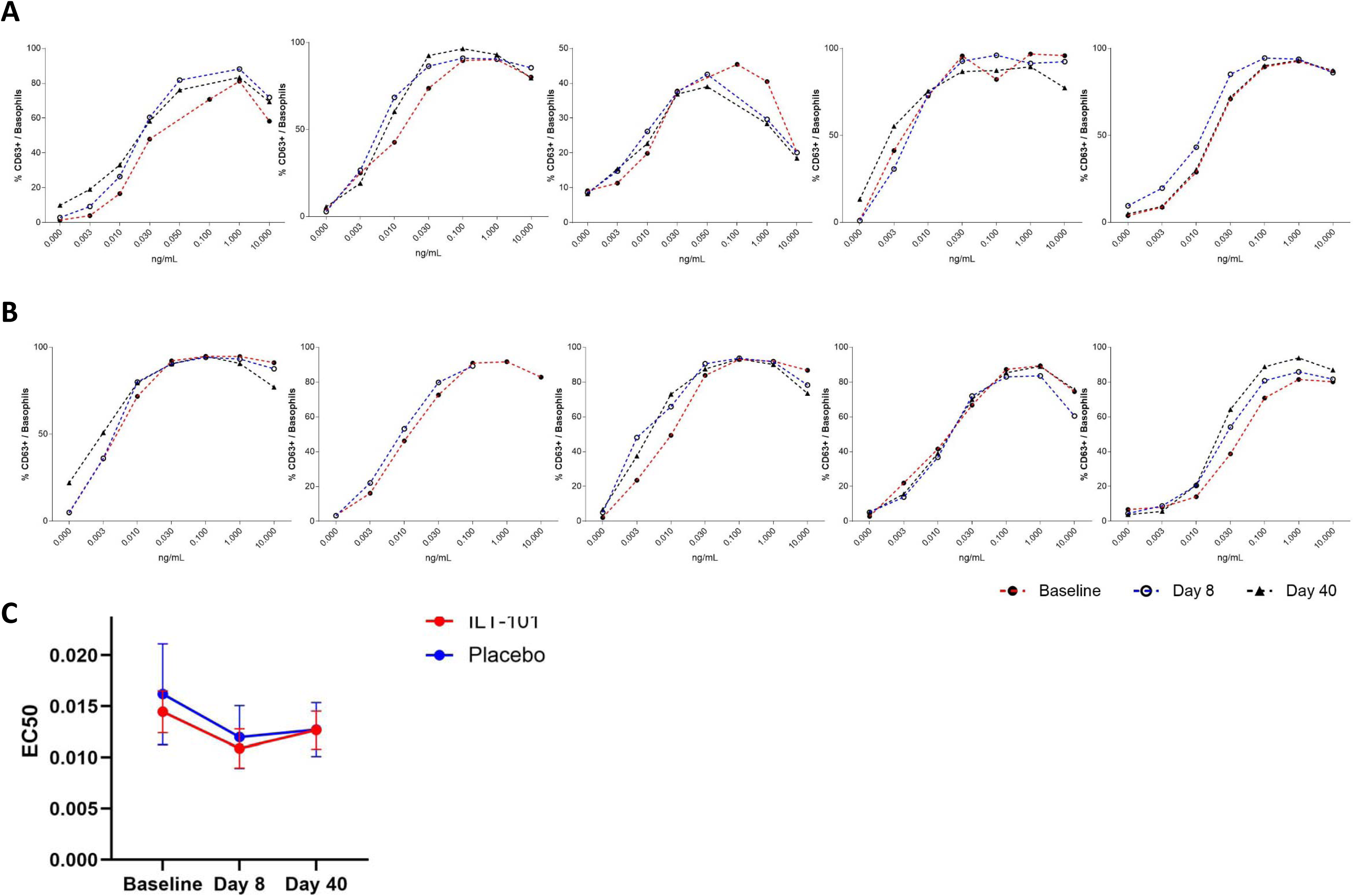
Changes in total and specific IgE and IgG4 serum concentration. Total IgE, allergen specific IgE and IgG4 were measured at baseline (day 1) and day 40 and are shown for each patient in ILT-101 treated group (in red) and in placebo group (in blue). Total IgE and specific IgE are expressed as kUI/L, total IgG4 as g/L and specific IgG4 as mg/L.

**Figure 6:** In vitro response to birch pollen of patients’ basophils. Dose response curves of basophil activation (%CD63+CD203c+basophils) with increasing doses birch pollen at baseline, day 8 and day 40 in **A)** ILT-101 treated patients (n=6) and **B)** in placebo patients (n=6); **C)** Representation of the calculated EC50 (mean± sd) in ILT-101 treated patients (in red) and in placebo patients (in blue)

## Discussion

No studies investigated the effects of IL-2_LD_ in allergy. In this study of birch tree pollen-induced allergic rhinitis, low dose of IL-2_LD_ induced time-dependent improvements in nasal symptoms assessed by TNSS as compared to baseline, but not relative to placebo. However, the rhinitis sign and symptoms had less clinical impact as revealed by the VAS patient-reported nasal questionnaire score. While VAS is a subjective scale assessing the global rhinitis, it is highly corelated to quality of life questionnaire that is frequently used in allergen immunotherapy studies^33,41,42^ . This indicates that the IL-2 effects for a short 6-week treatment are not sufficient to demonstrate a major effectiveness in a small number of patients. As within the IL-2 treated group there is a progressive time dependent increase in clinical improvements, this suggest that longer treatment periods could lead to significant clinical improvements.

The improvements in the rhinitis symptoms are associated with an IL-2_LD_-induced significant increase in the number and activation of Tregs, as observed in all other diseases tested^43^. Thus, our study reveals that Tregs from allergic patients have the same sensitivity to IL-2 activation as Tregs from healthy volunteers or patients with autoimmune diseases^43^. The short nature of the IL-2_LD_ treatment could not affect significantly the concentration of allergen-specific IgE, which remained unchanged in both treated and placebo groups, and it likewise did not affect the allergen threshold sensibility of basophils degranulation. This immunologic data supports that the increasing fitness of Tregs likely mediated the clinical improvements. This in turn indicates that the function of Tregs is not only the maintenance of tolerance to allergens, preventing allergy in healthy individuals, but also the mitigation of the allergic reaction whenever it occurs in allergic individuals.

The results of this study show that IL-2_LD_ therapy is well-tolerated, as has already been demonstrated in several other published studies on patients with different autoimmune and chronic inflammatory diseases. No serious adverse events related to treatment were reported, and the reported, mild or moderate, adverse events were those already widely known from IL-2 therapy ^44–47,37,40,48,49,38,43^. This is particularly important for further use of IL-2_LD_ in allergy. Indeed, there were initial concerns with using IL-2 in the treatment of autoimmune diseases because it could have stimulated the effector T cells that mediate the disease. These concerns were relieved after showing that at low doses, IL-2 specifically activates Tregs but not Teffs. This licensed many clinical trials of IL-2_LD_ that have reported a good safety profile and the universal effects of IL-2_LD_ in expanding and activating Tregs, without effects on Teffs and with no aggravation of diseases. These trials also reported a universal IL-2_LD_-induced eosinophilia that was later linked to the IL-2_LD_-induced stimulation of IL-5 production by ILC2^50^. This eosinophilia has legitimately cast concerns on the safety of investigating IL-2_LD_ in allergy, despite a strong rationale for doing so. While around 50% of our patients had concomitant asthma, we did not register any asthmatic manifestation after ILT-101 administration. Moreover, the design of our study in which there is a controlled allergen administration in an EEC allowed for accurate follow-up of the clinical manifestation of the allergic response. Spirometry evaluation performed every 20’ after the challenge and for 4 hours revealed an improvement in these respiratory functions in the IL-2_LD-_treated patients compared with placebo. Thus, our work relieves safety concerns of using IL-2_LD_ in allergy and licenses its further investigation, including in asthma.

Such investigation should involve treatment of longer duration that might also have an impact on some of the mechanisms of allergy, and notably the IgE production. Indeed, prophylactic IL-2 treatment expand Tregs and inhibit allergen-specific sensitization in experimental models^51^. As the stimulation of Tregs represents a novel target in allergy, it could be combined with treatment targeting other pathways. In this line, combination therapies with anti-IL-5 treatment have an obvious rationale. Finally, the ultimate goal for allergy treatment is the re-establishment of tolerance to the allergen. AIT has a clear but limited efficacy, which has furthermore been linked to the induction of Tregs. The addition of an IL-2 treatment to AIT might favour the development of allergen-specific Tregs and improve efficacy. Further studies are warranted to define the potential of Treg stimulation in the prevention and treatment of allergy.

## Data Availability

All data produced in the present study are available upon reasonable request to the authors

## Authors’ contribution to the current study

DK conceived and supervised the study.

MR, RL, EV, BB, AS, FdB and DK designed the study.

AG, FD, MB, ND performed the clinical follow-up of patients under the supervision of FdB.

MR supervised the immunomonitoring.

MR, and DK analysed the biological results.

DdB analysed the clinical results.

MR and DK wrote the first draft of the manuscript that was edited by all authors.

## Acknowledgement

This study was funded by the Programme Hospitalier de Recherche Clinique, N° P160936J . The Assistance Publique-Hôpitaux de Paris sponsored the study and ILTOO pharma provided the treatments. We thank the personnel of the Pitié-Salpêtrière Clinical Investigation Center for Biotherapies (CIC-BTi), Michèle Barbié, Natalie Féry, Catherine Ferrapie, Nadia Graffin, Aurélie Marc, Alexandra Roux and Fabien Pitoiset for their excellent assistance. We thank Dominique Batejat for supervising treatment provision.

