## Supplemental materials for "Low-dose interleukin-2 in birch pollen allergy: a phase-2 randomized double-blind placebo-controlled trial"

SUPPLEMENTARY APPENDIX
TABLE OF CONTENT
- Table S1: Inclusion/Exclusion criteria
- Table S2: Primary and secondary Outcomes
- Figure S1: Study design
- Figure S2: Eosinophils activation markers
- Supplementary Methods


Table S1: Inclusion/Exclusion criteria
PATIENTS
Main inclusion criteria
- Male or female aged [18-55] years
- Positive clinical history of seasonal allergic rhinitis to birch pollen for at least 2 consecutive pollinic seasons before inclusion and requiring medication intake with or without GINA 1  associated asthma, with documentation of sensitivity within 12 months before enrolment, by positive skin prick test (SPT) and by validated in vitro tests for specific Immunoglobulin E (IgE);
- Positive nasal response (TNSS?5) during baseline exposure 
- Negative urinary pregnancy test at screening visit for women of childbearing age;
- Ability to stay in the EEC for up to 4 hours, without any conditions or factors which could make this not possible
- Free, informed and written consent signed by the patient and the investigator, before any specific examination required by the study;
- Affiliation to a social security scheme (beneficiary or assigned)
- Negative SARS-CoV-2 test less than 96 hours prior to screening visit
Main non-inclusion criteria
- Asthma: GINA 2 to 5
- Eosinophilia >0.6.109/mL;
- Any history of anaphylactic reactions;
- Specific immunotherapy treatment at the moment, including Omalizumab; 
- Specific immunotherapy for birch-pollens within 3 previous years; 
- Use of systemic corticosteroid or others immunosuppressive treatment within previous 6 months;
- Moderate to severe allergic rhinoconjunctivitis with or without asthma due to grass pollen (according to ARIA)
- Significant rhinitis, or sinusitis, due to daily contact with other allergen causing symptoms that are expected to coincide with exposures, as assessed by the investigator
- Contraindications known to treatment with IL-2:
- Hypersensitivity to the active substance or to any of the excipients;
- Immunosuppressed patient;
- Psychotropic, hepatotoxic, cardiotoxic nephrotoxic, drugs;
- Other chronic diseases not clinically controlled;
- Signs of active infection requiring treatment;
- Previous history of organ transplantation.
- Heart failure (? grade II, class. NYHA), kidney failure (Cockroft <60 ml/min/1.73m2), liver failure (transaminase> 3N), pulmonary insufficiency (any grade).
- Leukocytes <3000 / mm3 lymphocytes <800 / mm3, platelets <80 000 / mm3, Haemoglobin < 10.0 g/dL or 6.2 mmol/L, red cell blood < 3.5 T/L;
- Chronic uncontrolled arterial hypertension (Systolic BP > 140 mmHg and/or Diastolic BP > 90 mmHg);
- Poor venous capital will forbid blood samples;
- Vaccination with attenuated live vaccine in the month before the inclusion or planned during the study;
- Surgery in the previous three months or anticipated under study;
- Participation in other interventional research with study drug in the previous month and during the study;
- Psychiatric illness or any other concomitant chronic illness or addiction that could interfere with the ability to meet the requirements of the protocol or provide informed consent;
- Presence or history of unhealed cancer for more than five years, presence or history of healed cancer for less than five years, except carcinoma in situ of the cervix or basal cell carcinoma;
- Pregnant or lactating women;
- Men and women of childbearing age without effective contraception during the treatment period;
- Patient with a measure of legal protection.

Table S2: Primary & secondary endpoint

ILT101
LS Mean
(95%CI)
Placebo
LS Mean
(95%CI)
ILT101 vs Placebo
p- value
Primary endpoint


Absolute change from baseline in TNSS AUC0-4h at day 40                 
-8.03 
(-12.90 ; -3.15)
-4.76 
(-10.06 ; 0.55)
0.3213
Secondary endpoint


Absolute change from baseline in Tregs % at Day 8
4.56 (3.04 ; 6.09)
0.27 (-0.39 ; 0.94)
0.0001
Absolute change from baseline in Tregs % at Day 40
2.85 (1.47 ; 4.22)
0.22 (-0.36 ; 0.79)
0.0018
Change from baseline in Nasal Response Intensity (VAS) AUC0-4h at day 40
-6.65 (-9.48 ; -3.81)
-3.35 (-8.27 ; 1.57)
0.2142
Change from baseline in FEV1P AUC0-4h at day 40 

11.78 (-4.70 ; 28.27)

-6.50 (-17.85 ; 4.84)
0.0427
Change from baseline in FEF25-75 AUC0-4h at day 40

0.76 (-0.58 ; 2.10)

-0.71 (-1.51 ; 0.08)
0.0384


Figure S1: study design


Figure S2: Eosinophil activation markers

Supplementary methods
EEC
ALYATEC EEC is a 65 m2 International Standards Organization (ISO) 8 clean room (Standard 14644-1) with continuous air circulation, containing 20 seats to accommodate up to 20 patients at 1 time, and with an anteroom and exit room. The EEC contains 10 particle counters spread equally throughout the room, and 5 glass fibber filters adjacent to seats to be used for Bet v 1 allergen testing by ELISA. Standardized, commercially available birch pollen extract prepared by a pharmacist is nebulized into the EEU through a specialized system that maintains consistent birch allergen particle counts and Bet v 1 ELISA dosing approximating 60 ng/m3 across the 20 seat positions of the EEC. Allergen particle plateau fall below the limit of detection within minutes of stopping the nebulization and Bet v 1 measurements are below the limit of quantification outside of the EEC (e.g., in the observation area and in the rest rooms). Other allergens have been tested in the EEC, observation room, and the rest rooms and are below the limit of quantification (e.g., house dust mite, cat), suggesting that there is no cross-contamination of allergens. During each exposure, the patients wear protective suits (TS Plus Microgard 2000) to ensure that no contaminant enters or exits the EEU. The cleaning procedure after each EEU exposure session includes the following: rinsing of the nebulization system, disinfectant of the floor, walls, the armchairs, and of all the small material stored in the EEU (Ecowipes, THX medical). Air qualification is performed by an independent company (Air Qualif) 4 times a year to test for the absence of bacteria and molds in the EEU. This facility enables the provision of a homogenous distribution of allergens for each participant. The airflow system, environmental conditions, and airborne particles are continuously monitored. Communication between participants inside the EEC and with medical supervisors was enabled using wireless telecommunication.

Flow cytometry
Blood samples were collected in EDTA tubes according to the planned protocol: absolute numbers of lymphocyte subsets and Tregs were monitored at each patient visit: day-1 (baseline) before treatment and during IL-2 administration at day 8 and day 40 of the follow-up. The last day follow up was 1 months post treatment at day 70. 
Blood subsets (CD3+, CD4+, CD8+ T lymphocytes, CD19+ B lymphocytes and CD3-CD56+ NK cells) counts (cells/?l) were established from fresh blood samples using CYTO-STAT tetraCHROME kits with Flowcount fluorescents beads and tetra CXP software with a FC500 cytometer (Beckman Coulter) according to manufacturer’s instructions. Foxp3 labeling was performed with custom designed dry tubes (duraclone from Beckman Coulter) containing CD3-FITC, CD25-PE, CD127-PE-Cy7, Foxp3-AF647, CD8-KRO, CD8-PB antibodies as described already described1. Total blood (60 µL) sampled with anticoagulant was mixed with 6 µL of Perfix-NC R1 buffer, vortexed immediately for 2-3 seconds and incubated for 15 min at room temperature in the dark. Perfix-NC R2 buffer (360 µL) was added and, after homogenization, a 360 µL aliquot was transferred to a Duraclone tube. After vortexing for 10 seconds, tubes were incubated for 60 min at room temperature in the dark. PBS 1X (3 mL) was added to the tubes, incubated for 5 min at room temperature in the dark before centrifugation for 6 min at 250g. The supernatant was removed to leave the pellet dried and the cells were resuspended in 3 mL of 1X Perfix-NC R3 buffer prior to another 6-min centrifugation at 250g. The pellet was dried and resuspended in 300 µL of 1X R3 buffer. Tubes were protected from light and stored at 4°C until the acquisition on a cytometer within the next 24 h. Acquisition was performed on a Navios cytometer (Beckman Coulter) maintained daily according to the manufacturer’s recommendations with Flow Check Pro and Flow Set Pro fluorospheres. Acquisitions were performed, respectively, using Navios software, and all analyses were done with Kaluza 1.3 software (Beckman Coulter)1.
Deep Immunophenotyping was performed as already described to study Eosinophils and Tregs activation using the following antibodies:  DR-FITC CD44-PE, CD69-ECD, CD244 PE-Cy5.5, CD14-PE-Cy7, CD123-APC, CD25-APC-AF700, CD16-APC-AF750, CD15-PB and CD45-KrO for eosinophils and CD127-FITC, GITR-PE, CD39-PE-Cy5.5, LAP-Percp-Cy5.5, CTLA-4 PE-Cy7, LAG3-ECD, FoxP3-A647, CD45RA-APC-AF750, Helios -PB, CD3-KrO, ICOS-BV610.  CD25-BV650, CD73-BV786, CD8-APC-AF700, CD4-BUV395. . Acquisition was performed using a LX CytoFLEX with Cytexpert Software and all analyses were done with Kaluza 2.0 software (Beckman Coulter)1
Basophil Activation Tests 
Basophil Activation Tests were performed using custom design kit Beckman Couter in which a proprietary dry reagent formulation technology  was used to prepare ready-to-use and room temperature stable reagents as already discribed2. Flow cytometry staining reagents (CD45-KrO (Clone J33), CD3-PC7 (Clone UCHT1), CRTH2-A647 (Clone BM16), CD203c-PE (Clone 97A6) and CD63-PB (CLB-Gran/12) all from Beckman Coulter, Marseille, France) alone or in combination with either anti-IgE (Clone E124.2.8, Beckman Coulter, Marseille, France) or Birch extract (BAGT3 antigen) at different concentration were dried together in the considered tubes (duraclone). Upon mixing and incubation 100µl of fresh blood at 37 °C for 15 min, erythrocytes were lysed and samples were analyzed by flow cytometry without further washing steps. Acquisition was performed using a LX CytoFLEX with Cytexpert Software and all analyses were done with Kaluza 2.0 software (Beckman Coulter). 
Basophils were gated as side scatter (SS) Low/CD45+/CD3?/CRTH2+ and both CD203c and CD63 were used to characterize basophil activation status following ex vivo activation in patients’ whole blood with increasing concentrations of allergen extract (0. -1000 pg/100µl) to generate a dose-response curve. Basophil responsiveness to birch was measured by the percentage of basophils positive for CD63 and CD203 surface expression in flow cytometry analysis and by the measured of EC50 (the allergen concentration required to achieve 50% of maximal basophil activation). A higher EC50 value indicates lower basophil responsiveness to allergen stimulation.
Total and specific IgE and IgG4 serum concentration 
Total IgE, allergen specific IgE and IgG4 were measured by ImmunoCAP™ corresponding reagents on a Phadia250 instrument (ThermoFisher Scientific, Upsalla, Sweden). Total IgG were measured using the IGG kit (Ref 446400) by on a IMMAGE instrument (Beckman Coulter). Total IgG4 were measured by an in-house method as described by Aucouturier et al3.
REFERENCES
1	Pitoiset F, Barbié M, Monneret G, et al. A standardized flow cytometry procedure for the monitoring of regulatory T cells in clinical trials. Cytometry B Clin Cytom 2018; published online Jan 6. DOI:10.1002/cyto.b.21622.
2	Arif-Lusson R, Agabriel C, Carsin A, et al. Streamlining basophil activation testing to enable assay miniaturization and automation of sample preparation. J Immunol Methods 2020; 481–482: 112793.
3	Aucouturier P, Danon F, Daveau M, et al. Measurement of serum IgG4 levels by a competitive immunoenzymatic assay with monoclonal antibodies. Journal of Immunological Methods 1984; 74: 151–62.
